# Next Generation Evidence: High-Precision Information Retrieval for Rapid Clinical Guideline Updates

**DOI:** 10.1101/2024.12.02.24318184

**Authors:** Florian Borchert, Paul Wullenweber, Annika Oeser, Nina Kreuzberger, Torsten Karge, Thomas Langer, Nicole Skoetz, Lothar H. Wieler, Matthieu-P. Schapranow, Bert Arnrich

## Abstract

Delays in translating new medical evidence into clinical practice hinder patient access to the best available treatments. Our data reveals an average delay of nine years from the initiation of human research to its adoption in clinical guidelines, with 1.7–3.0 years lost between trial publication and guideline updates. A substantial part of these delays stems from slow, manual processes in updating clinical guidelines, which rely on time-intensive evidence synthesis workflows. The Next Generation Evidence (NGE) system addresses this challenge by harnessing state-of-the-art biomedical Natural Language Processing (NLP) methods. This novel system integrates diverse evidence sources, such as clinical trial reports and digital guidelines, enabling automated, data-driven analyses of the time it takes for research findings to inform clinical practice. The NGE system accelerates guideline updates by employing precision-focused literature search filters tailored specifically for guideline maintenance. In benchmarking against two German oncology guidelines, these filters demonstrate exceptional precision in identifying pivotal publications for guideline updates. By streamlining evidence synthesis, NGE has the potential to deliver faster updates, improve guideline responsiveness, and enhance patient access to state-of-the-art treatments.

## 1 Introduction

The past years have witnessed remarkable advances in biomedical Natural Language Processing (NLP), significantly enhancing the ability to extract meaningful insights from unstructured sources of medical evidence, including clinical trial reports and clinical guidelines [1, 2]. While primary research publications have received considerable attention by the NLP community, the textual contents of clinical guidelines remain under-utilized, especially for languages other than English. Recent innovations in multilingual and domain-specific medical language models have greatly improved the viability of using data from international clinical guidelines in software systems and support the timely translation of clinical research into actionable recommendations for healthcare decision-making [3, 4].

The translation of new evidence into clinical practice is currently impeded by multiple delays throughout the process [5]. A substantial factor contributing to these time lags is the inherent complexity of clinical trials, which require extensive time for ensuring safety, efficacy, and robust data collection [6]. Yet, the volume of published research results in the primary medical literature is still so large, that another bottleneck becomes *evidence synthesis*, i.e., a summary of the body of evidence with a critical appraisal of its quality and impact for clinical practice [7]. Approximately one million articles dealing with clinical trials are currently indexed by PubMed, more than 30 thousand of them have been added in 2023 alone (about 82 per day)^1^. In effect, incorporating all available evidence into evidence synthesis workflows for guideline development becomes increasingly time-consuming.

In this work, we focus on the particular delay induced by current update protocols for clinical guidelines, i.e., the time it takes to incorporate successfully published research results into guideline recommendations. These protocols vary across guideline groups, but they usually involve a formulation of key questions using the Population–Intervention–Comparison–Outcome (PICO) framework, a systematic literature search, data extraction, assessment of the robustness of the underlying evidence, as well as procedures to arrive at recommendations through evidence-to-decision (EtD) frameworks and structured consensus-finding processes [8, 9]. For literature retrieval in these projects, most guideline developers follow similar approaches, i.e., typically a Boolean search in literature databases such as PubMed [10]. These searches tend to aim for near-perfect *recall*, while suffering from notoriously low levels of *precision*, i.e., most search results are irrelevant [11]. Common search queries may return thousands of results, which need to be reviewed manually by human experts through screening of title, abstract, and full-text [12]. Recently, the concept of *living guidelines* gained increased attention: these are supposed to be updated as soon as new evidence becomes available, ideally on the level of individual recommendations [13]. A natural implementation of such a surveillance strategy would be the regular application of an existing search query to the stream of newly published literature [14]. However, this would not alter the overall screening burden incurred by low retrieval precision of existing search strategies.

Assuming that comprehensive literature reviews within the scope of a guideline topic are performed at regular intervals, all relevant publications should be covered by such a review at some point in time. Thus, a complementary search strategy aiming for high precision instead of perfect recall can focus on *signal publications*, i.e., publications likely to significantly influence the conclusions or recommendations of a review or guideline. Shekelle et al. [15] describe such an approach, based on “limited literature searches and expert opinion” in the context of systematic reviews. The American Society of Clinical Oncology (ASCO), has adopted a similar strategy for updating oncology guidelines, relying on “targeted literature searching and the expertise of ASCO guideline panel members” [16]. For example, ASCO has recently issued a rapid update of the non-small-cell lung cancer guideline based on the results of a single phase III clinical trial [17, 18].

Our work presents an innovative, data-driven approach to address challenges posed by intermittent updates to clinical guidelines. We propose to integrate diverse sources of primary and synthesized evidence automatically. In the remainder of the work, we share details about our developed *Next Generation Evidence* (NGE) system, illustrated in Fig. 1, which is leveraging recent innovations in multilingual medical NLP research. Our harmonized database ensures semantic interoperability by mapping all relevant information to concepts from the Unified Medical Language System (UMLS) [19]. Currently, we incorporate the following medical evidence sources:

- Digital clinical guidelines, which are input for various NLP components for structured information extraction developed in the context of the GGPOnc and xMEN projects [20–22],
- Reports of randomized controlled trials (RCTs) in PubMed, from which we automatically extract information related to PICO elements, inspired by the Trialstreamer [23] system,
- All registered clinical trials in ClinicalTrials.gov, which are accessed through the AACT database [24], as well as
- Curated assertions from CIViC, as a precision oncology knowledge base (POKB) for the clinical classification of cancer variants that is widely used by translational oncologists [25, 26]

**Figure 1.**
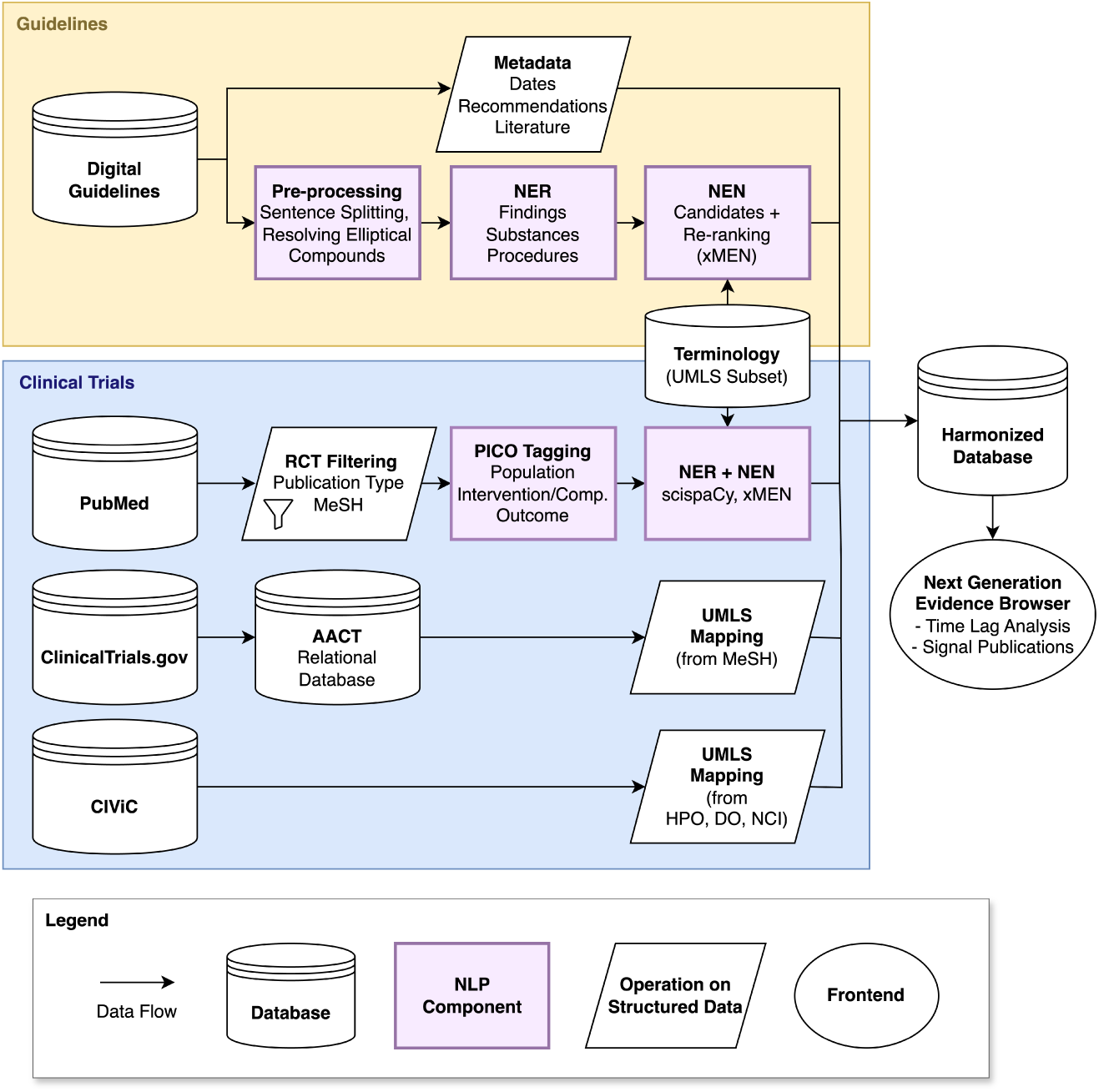
Integration of different sources of medical evidence into a harmonized database. We incorporate the contents of clinical guidelines, clinical trial reports in PubMed, registered clinical trials in ClinicalTrials.gov, as well as assertions from CIViC, a widely used knowledge base (KB) for precision oncology. As both guidelines and trial reports consist of mostly unstructured text content, we apply recently developed NLP components to extract structured data from these sources. Details on each component are provided in Tab. 1

**Table 1.** Data sources and software components for their integration. For each component, we indicate whether the component uses NLP and, if so, which kind of model.

| Source | Comp. | Description | NLP Model |
| --- | --- | --- | --- |
| Clinical Guidelines | Metadata | Extraction of structured metadata from a digital guideline repository | – |
|  | Pre-processing | Syntactic pre-processing (e.g., sentence-splitting); replacing elliptical coordinated compound noun phrases (“chemo- and radiotherapy”) with their expanded form (“chemotherapy and radiotherapy”) | Encoder–decoder model (mT5) trained with >3K manually annotated sentences in GGPOnc 2.0 [21, 46] |
|  | NER | Named entity recognition for findings, substances, and procedures within clinical guidelines | Nested NER model initialized from MEDBERT.DE and trained with >200K long, fine-grained entity annotations in GGPOnc 2.0 [20, 47] |
|  | NEN | Named entity normalization of mentions to UMLS concepts | xMEN candidate generation with a knowledge base (KB) initialized from a UMLS subset, followed by re-ranking [22] |
| PUBMED | RCT Filtering | Identification of RCTs in MEDLINE based on publication types and MESH terms | – |
|  | PICO Tagging | Identification of all PICO spans within MEDLINE abstracts | BioELECTRA model fine-tuned for PICO extraction on the EBM-NLP dataset [48, 49] |
|  | NER + NEN | Identification of all named entities within PICO spans and normalization to UMLS concepts | SCISPACY NER model trained on MEDMENTIONS, SCISPACY entity linker adapted to a custom UMLS subset [50, 51] |
| Clinical-Trials.gov (via AACT) | UMLS Mapping | Mapping of already normalized conditions and interventions (MeSH terms) to UMLS concepts | – |
| CIViC | UMLS Mapping | Mapping of already normalized diseases (DO), phenotypes (HPO), and therapies (NCI thesaurus) to UMLS concepts | – |

Moreover, we provide a user-friendly web application to interact with the database, which is available online: https://we.analyzegenomes.com/nge/. We believe that the system can be useful for numerous stakeholders. Amongst others, guideline developers can use it a) to implement targeted signal search strategies for clinical trial publications, e.g., for entirely new treatment options, b) to quality-check the results of traditional searches, but also c) to support existing (consensus-based) recommendations with more solid evidence. Furthermore, our tool can be used by guideline users to identify newly published evidence that might affect the interpretation of current recommendations prior to a guideline update [27].

We evaluate our system experimentally as follows. First, we perform an analysis across the integrated data sources with the goal to automatically estimate the time it takes to translate research on new treatments for cancer patients from the first clinical trials with human subjects to recommendations in oncology guidelines. Second, we show how the system can be used to identify signal publications, which might be relevant for prospective guideline updates. To avoid an unreasonable increase in the screening workload induced by existing search strategies (which our system complements rather than replaces), our approach aims for maximal precision, i.e., any retrieved publication should be relevant for guideline developers and users with high probability. Two recent updates to German oncology guidelines are used for evaluating retrieval performance in a real-world setting: (1) *oesophageal cancer* and (2) *Hodgkin lymphoma*. The data from these updates enables an assessment of precision and recall in comparison with established guideline update protocols.

The remainder of this work is structured as follows: We review related work on existing approaches for time-lag analysis and literature retrieval for medical evidence synthesis in Section 2. We describe the components of our NGE system and the experimental setup for its validation in Section 3. In Section 4, we describe the harmonized database and experimental evaluation of the system. Our work concludes with a discussion in Section 5 followed by an outlook in Section 6.

## 2 Related Work

The estimation of time lags in translation from research into clinical practice has received considerable attention in the past. In prior research, this question was approached using selected case studies across different medical fields and types of interventions based on a manual review of publications [28–30]. As a prominent example, Hanney et al. [5] performed such an analysis across 11 calibration points in clinical research. First, the authors reviewed earlier studies on time lags in research translation, which found an average time span of 17 years from basic research to inclusion in guidelines. In their assessment, the authors find widely varying time lags from *discovery* (basic research) to *implementation*, ranging from 18 years (early interventions for schizophrenia) to 54 years (smoking reduction). Instead of relying on case studies, our work attempts a data-driven automation of such an analysis for a subset of this timeline, ranging from the beginning of research in humans (phase I clinical trials) until recommendation in a clinical guideline. In particular, we aim to assess the delay induced by the time it takes to perform a guideline update after results have been successfully published in the corresponding primary research literature, usually a medical journal.

There are several procedural and organizational challenges in clinical guideline development [9, 31]. Delays arise from the iterative steps required to identify and prioritize key clinical (PICO) questions, conducting literature reviews, assessing the robustness of the underlying evidence, and arriving at recommendations in more or less structured ways, e.g., through the GRADE EtD framework [32]. Further complexities arise from logistical aspects, e.g., team coordination, funding constraints, managing conflicts of interest, approval processes by guideline organizations, and quality control according to international standards such as the Agree II tool [33].

Among the aforementioned potential bottlenecks, previous work has pointed out the substantial delay that is induced by the excessive time it takes to conduct comprehensive literature reviews as the basis for evidence-based guidelines. Conducting a systematic review can take several years, whilst at the same time, reviews are prone to become outdated as new evidence is published [34, 35]. There is a wide range of techniques for automating parts of this process, e.g., through classification of publications according to inclusion criteria, risk-of-bias assessment, or extraction of structured data into evidence tables [36, 37]. Recent related work implementing such approaches are RobotReviewer [38] or Trialstreamer [23]. However, these research prototypes are not yet widely adopted in practice due to their lack of validation.

Although our NGE system does not seek to automate systematic reviews but rather complement them through more targeted search strategies, work in systematic review automation has led to valuable contributions. For instance, our pipeline for integrating RCT reports resembles the Trialstreamer workflow [23], but has been adapted in various ways, e.g., by relying more extensively on (automatically) assigned metadata, as well as the modification of information extraction components for PICO tagging, Named Entity Recognition (NER) and Named Entity Normalization (NEN), with more recent and customized implementations.

Improving precision for medical literature search has been studied extensively in the last decades [39]. Specifically, a multitude of approaches was proposed to filter and rank collections of clinical trial reports and other medical publications. PubMed, probably the most widely used medical search engine, allows fine-grained filtering based on text matches, MeSH terms, and other metadata. Recently, the National Library of Medicine (NLM) has adopted deep-learning-based automated indexing approaches using the Medical Text Indexer-NeXt Generation (MTIX) system, making such metadata available in a more timely manner [40]. Furthermore, PubMed has introduced a *Best Match* option, considering different metrics and the user query for ranking search results [41]. Even more advanced search options are provided by thirdparty systems and meta-databases, such as the Trip database [42] and other research prototypes [43–45]. To our knowledge, the NGE system is the first of its kind that contextualizes literature searches with the current state of recommended clinical practice, e.g., by focusing on interventions, which are currently not mentioned or recommended in a given clinical guideline.

## 3 Materials and Methods

This section describes our incorporated methodology for obtaining an integrated database of clinical evidence, a software application for interacting with these data, and the experimental setup for the evaluation of our system.

### 3.1 Data Integration

The NGE database builds upon periodically replicated copies of the underlying, heterogeneous data sources, i.e., clinical guidelines and various sources of primary evidence, such as the description and results of clinical trials. The data from each source is processed by individual Extract–Transform–Load (ETL) processes [52]. The ETL results are stored as a materialized version in a relational database. Fig. 1 and Tab. 1 provide an overview of the involved ETL components for each of our currently integrated data sources; a detailed description is given in Supplementary File 1. Many data sources provide mostly unstructured data (clinical guidelines, PubMed abstracts). Therefore, we employ a variety of NLP components for NER and NEN. Since international guidelines are published in many national languages, these data require language-specific NLP tools. Presently, we focus on German oncology guidelines that we obtain from a Content Management System (CMS) maintained by the German Guideline Program in Oncology (GGPO) [53]. Nonetheless, our system can be extended to incorporate also other sources, e.g., the Magic app [54] or adapted to incorporate international standards, e.g., CPG-on-FHIR [55]. For the unstructured portions of the guidelines (recommendations and background texts), we leverage NER models developed in the context of the GGPOnc project, as well as the xMEN toolkit for cross-lingual entity normalization to map these entities to Concept Unique Identifiers (CUIs) from a custom, task-specific UMLS subset [20, 22].

Mapping to UMLS CUIs ensures a high degree of semantic interoperability across overlapping items from integrated data sources. Moreover, evidence in all sources can be linked to one or more *population* and *intervention* attributes, although with different naming conventions. This mapping to populations and interventions is usually straightforward. However, the following design decisions were taken. First, *clinical drugs* and *therapeutic procedures* are considered as interventions in guidelines, based on the fine-grained named entity classes identified by the GGPOnc NER tagger [20]. Second, all *phenotypes* and *diseases* are considered as populations for CIViC.

### 3.2 The NGE Browser

The key use case of the NGE database is to query its content based on a clinical question, formulated with respect to population or intervention concepts, and to filter the results according to various criteria, such as publication timestamps. The database is accessible through both a REST API and the NGE browser, a user-friendly web application, which allows researchers, guideline developers, clinical practitioners, and other potential users of the system to easily interact with the data.

#### 3.2.1 Searching for Clinical Trials

Fig. 2 depicts the search view of our NGE browser, which allows retrieving RCTs based on a particular population, and filtered according to different selection criteria such as publication dates or clinical trial phases. The user has to select at least a population, here given by a guideline topic. When using the system without the graphical user interface but through its exposed REST API, it can also be queried with custom sets of UMLS concepts, e.g., to find evidence for a specific subpopulation. The result is a list of RCTs from different sources, i.e., PubMed articles retrieved through Medline or CIViC, or clinical trials registered at ClinicalTrials.gov. To guide the user to the relevant search results, the tool highlights extracted interventions using color codes based on their occurrence within the selected guideline that is related to the selected publication. The search view offers various selection criteria to filter the result set according to different requirements a user may have. The default values are tailored to a prospective scenario, i.e., targeted for users, who wish to identify new, potentially practice-changing evidence with respect to an existing clinical guideline. A detailed description of the available filters is provided in Supplementary File 2. In the following, we focus on two main innovative features enabled by our employed NLP components.

**Figure 2.**
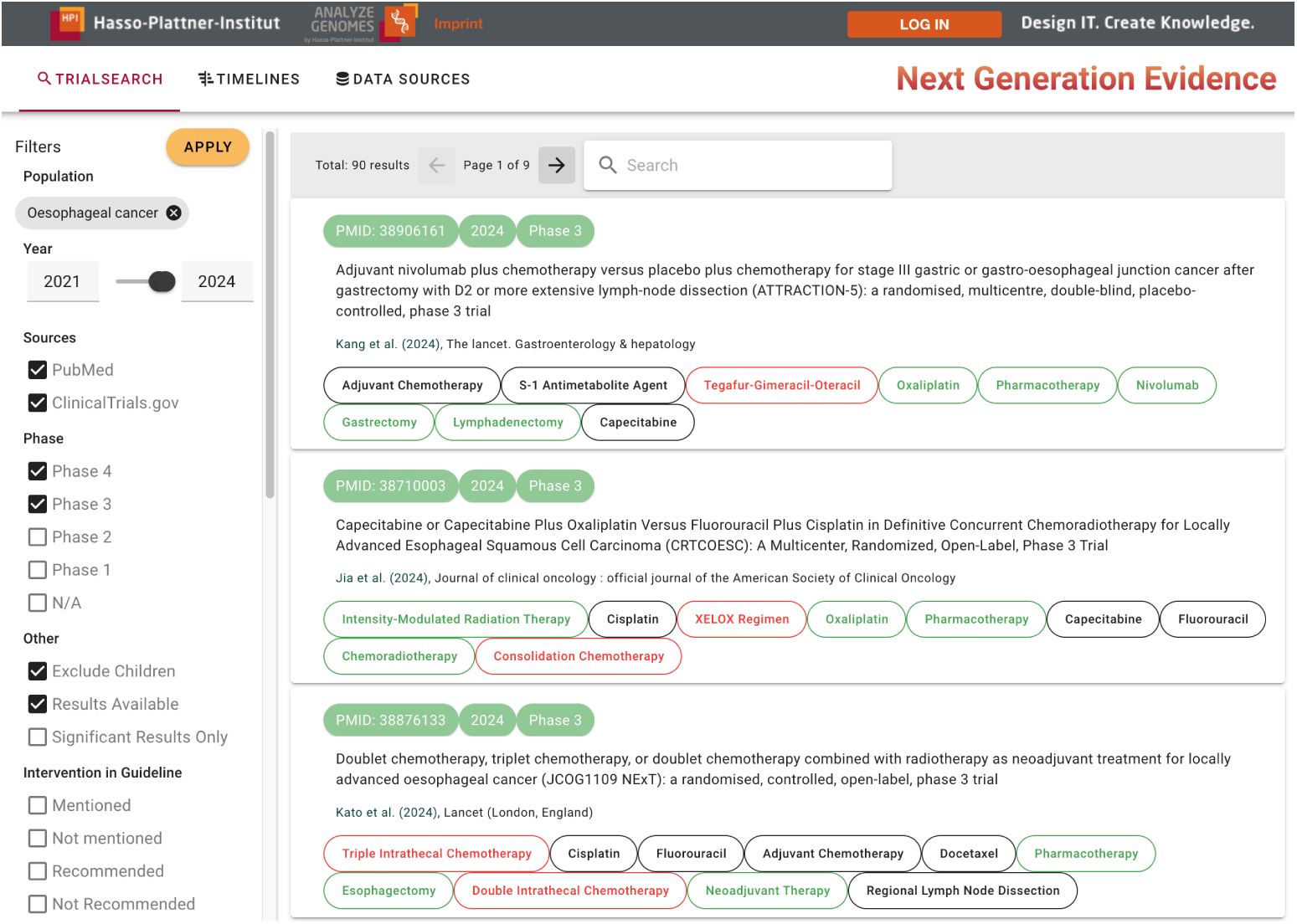
User interface for searching the NGE database for trials by population in combination with various filtering criteria. For all results, the system lists the identifier (PubMed or ClinicalTrials.gov ID), publication date, phase (I–IV), and title. For PubMed results, bibliographic information (authors and journals) is included. Extracted interventions are color-coded based on their occurrence in the guideline corresponding to the selected publication: black ones are already mentioned (anywhere) in the guideline, green ones are mentioned within recommendations, red ones are mentioned nowhere.

##### Interventions

Novel selection criteria stem from contextualizing the search with the current state of recommended clinical practice represented by the guidelines in the database. As highlighted in Fig. 2, interventions mentioned in a clinical trial can have different relationships to a guideline: they might be mentioned a) anywhere in the guideline, b) inside a recommendation, or c) not mentioned at all. We hypothesize that interventions that are not yet recommended or mentioned otherwise provide particularly strong update signals for an existing guideline.

##### Significant Results

We further suspect that the results from RCTs might be of particular interest when they report a significant improvement of some outcome of interest, especially for new interventions. Therefore, a flag is included to filter trials based on the statistical significance of their findings. In ClinicalTrials.gov, this information is often available as part of the structured results: here, we consider any RCT with a change in outcome associated with a *p* value lower than 0.05 as significant. For RCT reports in PubMed, the required details are obtained from the free-text abstract using a binary text classifier. Our employed classifier was obtained by fine-tuning a PubMedBERT model with annotations derived from the Evidence Inference 2.0 dataset [56, 57]. This feature is disabled by default because it is considered as highly experimental. More details can be found provided in Supplementary File 2.

#### 3.2.2 Visualizing Timelines

Fig. 3 depicts the timeline view in our NGE browser, which provides a temporal perspective of the current state of clinical research on a particular intervention in the context of current and previous guideline versions. The example shows the timeline view for the drug *Ipilimumab* for the clinical indication oesophageal cancer. Results of two phase III RCTs have been published just before the literature search for the latest update (version 4.0) has been performed. As a result, these were subject to the screening and data extraction phase of the review process and included in the guideline (green stars in the figure). However, shortly after finishing the search, two additional reports of phase III RCTs were published. Moreover, new data (subgroup analyses) for the two RCTs that were included were published in the meantime.

**Figure 3.**
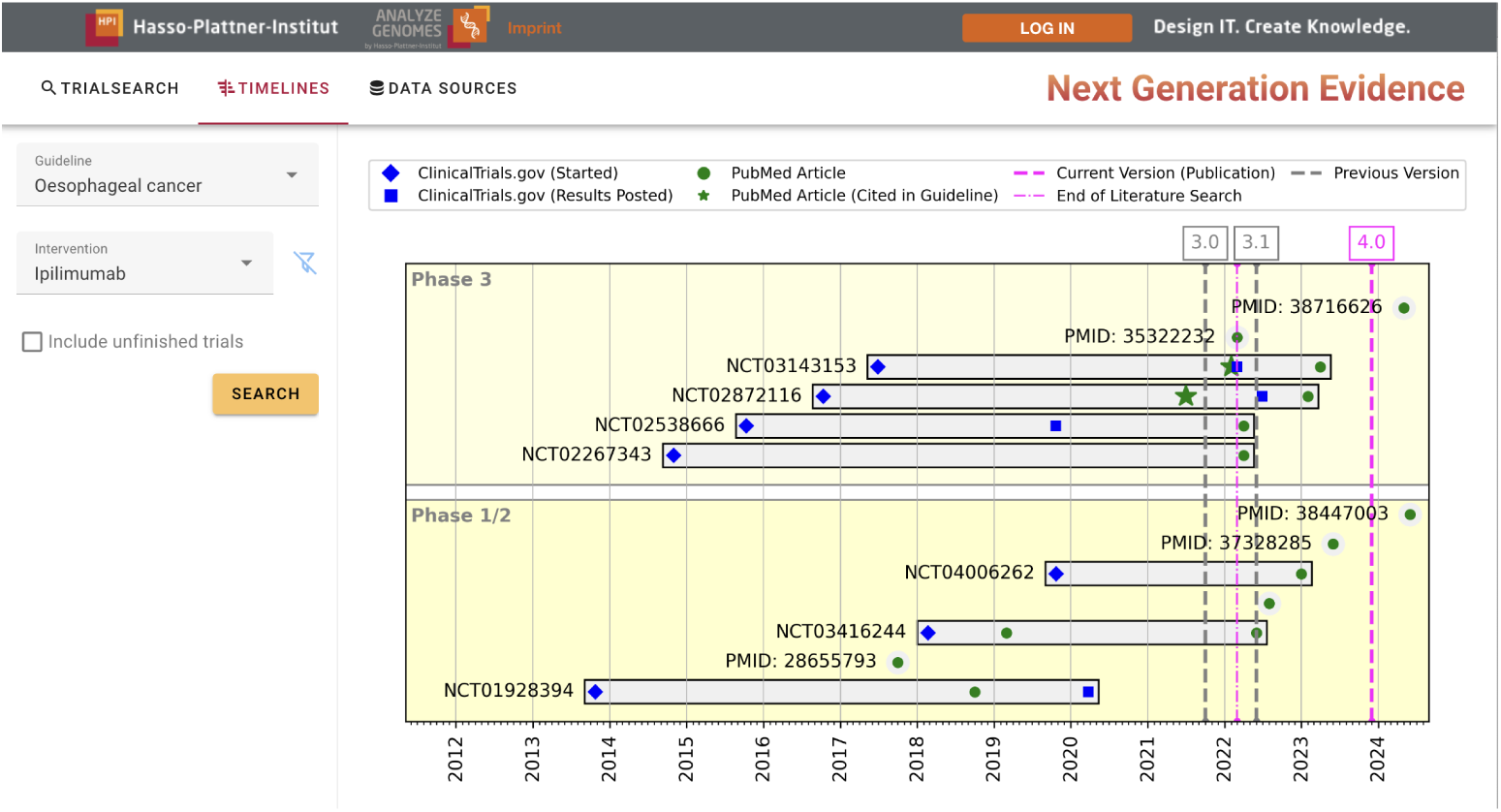
The timeline view groups registered clinical trials from ClinicalTrials.gov in terms of their start date, result publication data, and potential PubMed articles referencing this data on a horizontal timeline. Update intervals for the corresponding guideline are included as vertical lines.

### 3.3 Evaluation Datasets

#### 3.3.1 Time Lag Analysis

To evaluate time lags in translation from primary research to clinical guideline recommendations, we consider as input data all updates to the GGPO oncology guidelines for the years 2022–2024, i.e., between the GGPOnc releases 2.0 and 2.3 [20]. 12 guidelines (out of 34 maintained by the GGPO) received an update during the considered time frame, and a few have been updated multiple times (15 updates in total, including minor updates). Using the UMLS-normalized entity mentions and recommendation metadata, we identify new interventions, which have been recommended for the first time in any of the updated guidelines within the given time frame. In addition, the following filtering steps are applied, to ensure that the data is of sufficiently high quality for further analysis:

1. NEN confidence after re-ranking of at least 0.1,
2. Exclusion of generic interventions such as “chemotherapy”,
3. CUI belongs to the UMLS semantic network hierarchy “Pharmacologic Substance” (TUI: T121) or “Therapeutic or Preventive Procedure” (T061), and
4. At least one clinical trial with this intervention can be found in our NGE system.

Step 1 excluded many non-pharmacological interventions, as these are more challenging to normalize with high confidence. Step 4 excludes some genuine interventions such as “meditation-based stress reduction” or “Yoga” for endometrial cancer, where the current guideline recommendation is based on the cross-sectional guideline on complementary medicine, rather than individual trials for the particular combination of intervention and population.

#### 3.3.2 Guideline Updates

The utility of the NGE system for targeted literature searches is evaluated using two datasets of published evidence, screened by human experts for recent guideline updates: (a) the recently completed update of the German *oesophageal cancer* guideline (from version 3.1 to version 4.0) and (b) the ongoing update of the *Hodgkin lymphoma* guideline (from version 3.2 to version 4.0). These datasets cover the inclusion and exclusion decisions during literature screening and were generously provided by the respective guideline working groups as an export from their currently used literature management tools. The key characteristics of the two cleaned evaluation datasets are presented in Tab. 2.

**Table 2.** Constructing two evaluation datasets for guideline updates through different steps of the literature screening process (starting after title–abstract screening for Hodgkin lymphoma)

|  | (a) Oesophageal<br>Cancer | (b) Hodgkin<br>Lymphoma |
| --- | --- | --- |
| Minimum Publication Date | 09/01/2019 | 01/01/2016 |
| Search Date | 03/04/2022 | 06/01/2023 |
| Screened | 3,147 | – |
| – Duplicates | 139 | – |
| = Unique references | 3,008 | – |
| – *title–abstract excluded* | 2,741 | – |
| = title–abstract included | 267 | 168 |
| – *Full-text excluded* | 195 | 105 |
| + *Excluded, but already in guideline* | 9 | 2 |
| = Included (Evaluation) | 81 | 65 |
| <sup>⊆</sup> RCTs included | 26 | 25 |
| + Manual review | 9 | 6 |
| + Retrieved, already in guideline | – | 9 |
| = **RCTs included (final)** | **35** | **40** |
| <sup>⊆</sup> Other included | 55 | 40 |
| <sup>⊆</sup> *Excluded (Evaluation)* | 2,927 | 103 |
| <sup>⊆</sup> *RCTs excluded* | 290 | 24 |
| + *Manual review* | 6 | 11 |
| = **RCTs excluded (final)** | **296** | **35** |
| <sup>⊆</sup> Other excluded | 2,637 | 79 |

For the oesophageal cancer guideline update, the complete full-text and abstract screening decisions were provided. Out of 3,147 total references in the time frame from January 2019 to April 2022, 139 were duplicates retrieved from different sources (e.g., PubMed or the Cochrane database). Another 2,741 references were excluded during title–abstract screening, and 195 additional ones after full-text screening. However, nine excluded references were already cited in a previous guideline version, which is possible due to overlaps of the search time frame with previous minor updates (version 3.1). If these references were retrieved by the NGE system, they would (arguably) be regarded as relevant; therefore, we consider them in the evaluation. A total of 81 references were included, but since the system is designed to retrieve only RCTs, only the subset of 26 RCTs is considered for evaluation (line “RCTs included” in Tab. 2). Out of the 2,927 excluded references, 290 were RCTs (“RCTs excluded”). The search retrieved additional results that were included in the datasets (more details in Section 4.3.1). 15 additional references were considered for oesophageal cancer following another manual review (9 included, 6 excluded).

For Hodgkin lymphoma, the screening period overlapped with the screening period of the former guideline update. Hence, the NGE system retrieves additional results that were already incorporated in the previous guideline version; they were manually marked as included. These results constitute the final ground-truth for system evaluation (lines “RCTs included / excluded (final)”). For the Hodgkin lymphoma dataset, references excluded during title–abstract screening are not available. Therefore, this dataset contains only 168 references, which were subject to full-text screening. Out of these, 105 were excluded, and two were added as being already cited in the current Hodgkin lymphoma guideline. From the remaining 65 references, 25 were RCTs, which were complemented by manually reviewed references. Since the results from title– abstract exclusion are missing, only 35 excluded RCTs are available for the evaluation of the system regarding the Hodgkin lymphoma update.

### 3.4 Code Availability

The source code to create a local instance of the NGE system is available as opensource software on GitHub: https://github.com/hpi-dhc/nge_db/. A prototype of our NGE web application is online available at: https://we.analyzegenomes.com/nge/.

## 4 Results

In this section, we give an overview of the data integrated within the NGE database. Furthermore, we show how this data is used to a) estimate time lags in research translation and to b) retrieve signal publications for guideline updates.

### 4.1 Database Statistics

Tab. 3 shows the total number of documents for each source, as well as the total and unique numbers of population and intervention concepts. The seed CUIs for the 34 guideline topics are expanded to more than 17K population CUIs. Although most of them are unique, there is a certain degree of overlap in the subtrees descending from the root population concepts (the UMLS hierarchy allows a concept to have multiple direct ancestors). As an example, cancers of the *oesophagogastric junction* are partially covered by both the guideline for *gastric cancer* and the guideline for *oesophageal cancer*. The statistics also indicate that RCT reports and registered trials cover a much higher number of populations and interventions than guidelines and CIViC because the integrated clinical trial data is not limited to cancer patients. The relative number of unique CUIs in PubMed is much lower: the most frequently occurring CUIs belong to very general concepts, e.g., “patient”, “women”, “adult”, “neoplasm” (populations), or “treatment”, “placebo”, “administration”, “antineoplastic agent” (interventions). The numbers of unique concepts in ClinicalTrials.gov and CIViC are particularly low; presumably, because codes have been assigned by human curators based on controlled vocabularies rather than being automatically extracted from natural language text.

**Table 3.**
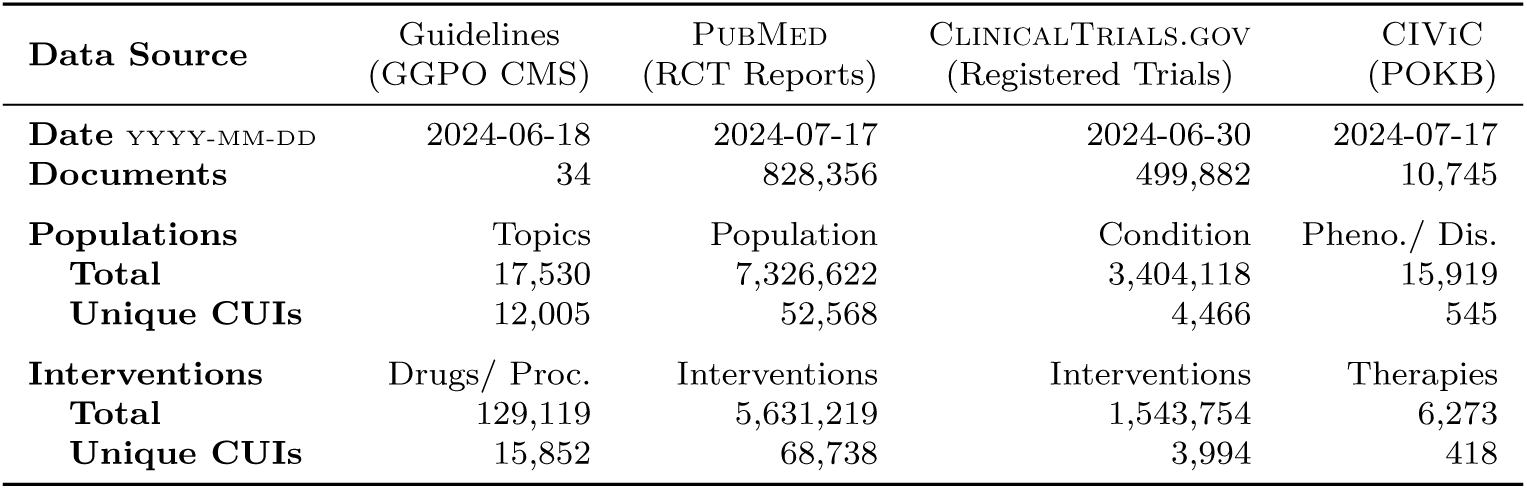
Overview of harmonized information for all integrated data sources. All results refer to data imported on July 17th, 2024, which was used for our evaluation. Data in the production system differs as it is regularly updated.

### 4.2 Time Lags in Research Translation

In total, 22 new interventions across oncology guideline updates could be identified from 2022 to 2024. All of these were manually validated as referring to new interventions in the focus of the respective guideline update. A detailed list of interventions and corresponding guideline updates is provided in Supplementary File 3. Fig. 4 shows a box plot of the distribution of time lags for all these interventions. The average time from the start of the first human trial to inclusion in a guideline recommendation is approximately nine years, which aligns with an estimate of eight to ten years by Subbiah [6]. As some guideline updates are based on results of phase I/II clinical trials, interventions with and without phase III trials are shown separately. When a guideline recommendation is based on the results of a phase I or II clinical trial, it takes an average of three years from publication to guideline recommendation. In contrast, this time span is reduced to 1.7 years for recommendations based on results of phase III clinical trials; presumably, because results from a large phase III trial might be a strong motivation for updating a guideline in the first place.

**Figure 4.**
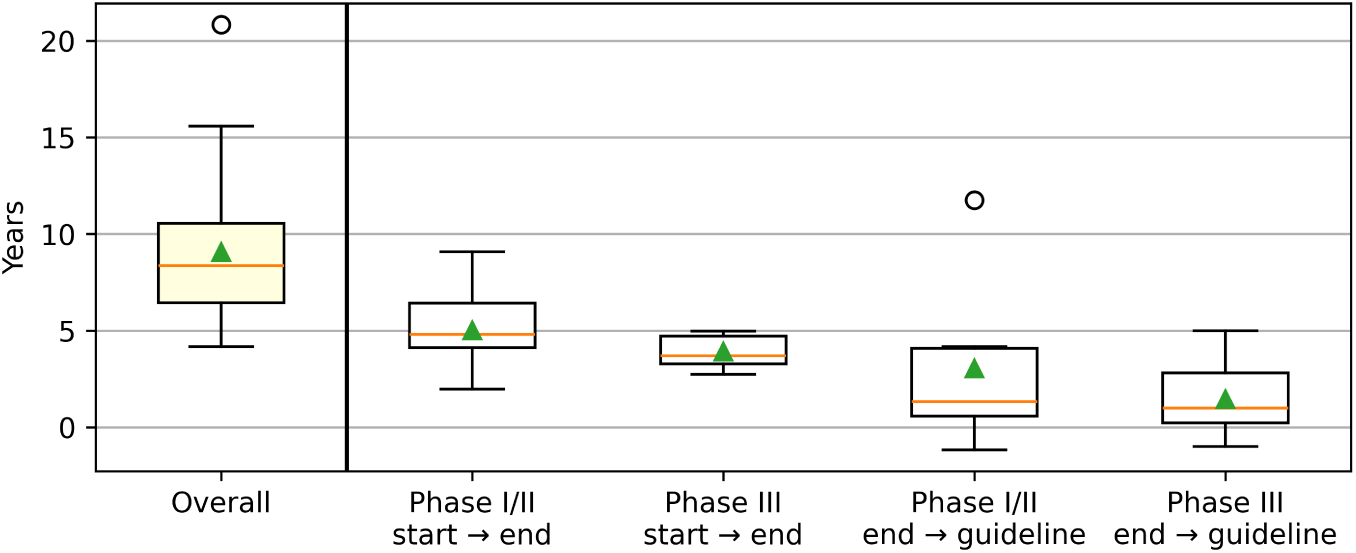
Box plot showing the distribution of time lags across all newly recommended interventions (n=22). The horizontal line within each box represents the median, the upper triangle the mean.

The standard deviation on all reported values is relatively large. The most visible outlier refers to one case, when a clinical guideline included results of a trial almost *12 years* after its publication. This contrasts with a few cases where a recommendation has been made even *before* the results of an ongoing clinical trial were published (negative values on the y-axis). Moreover, the visualized time frames refer to *publication* of results. Hence, phase III trials are frequently started before phase I/II results are available in the literature, potentially owing to the (well-known) delays in academic publishing. A qualitative description of example data points from Fig. 4 is provided in Supplementary File 3.

### 4.3 Retrieval of Signal Publications

To reduce the delay between publication of clinical trial results and inclusion in guidelines, the NGE system can be used to implement a targeted literature search approach. Thus, adding increasingly strict filter combinations allows users to balance between desired levels of precision and recall.

#### 4.3.1 Previously Unscreened Results

As described in Section 3.3.2, there are instances of results retrieved by the NGE system, which were not included in the initial screening at all. Such references were sent to the two groups that conducted the literature screening, asking them for additional feedback on the relevance of these results. However, only RCTs after phase II were subject to manual review to limit the workload because these were supposedly more likely to be relevant. For these references, human subject-matter experts deemed nine out of 15 results (60.0 %) potentially relevant for oesophageal cancer and six out of 17 results (35.3 %) for Hodgkin lymphoma. The assigned inclusion and exclusion reasons for these references are shown in Tab. 4.

**Table 4.** Inclusion and exclusion reasons for manually reviewed references.

| Manual Review | (a) Oesophageal Cancer | (15) | (b) Hodgkin Lymphoma | (17) |
| --- | --- | --- | --- | --- |
| Included | Potentially relevant | (3) | Potentially relevant | (6) |
|  | Drug not in PICO search | (6) |  |  |
| Excluded | Wrong population | (1) | Wrong population | (5) |
|  | Wrong publication type | (1) | Wrong publication type | (6) |
|  | Not in scope of update | (4) |  |  |

For Hodgkin lymphoma, four out of six relevant references (66.7 %) were actually part of the (temporally overlapping) literature search for the previous guideline version, but ultimately not considered by the guideline expert panel. Two references were erroneously excluded during title–abstract screening, the data of which is missing from the evaluation dataset for Hodgkin lymphoma. Regarding the oesophageal cancer guideline, where complete data is available, six references were not found originally, as the investigated drug was not explicitly part of the PICO (Boolean) search string. Regarding irrelevant results, the most common reason was a mismatch between the population and the scope of the guideline or the particular questions for the update. For instance, three results for Hodgkin lymphoma concerned children, which are out of scope of the guideline. Moreover, some results were indexed as RCTs in PubMed, but were, in fact, other publication types, e.g., secondary analysis of RCT data. The references that underwent a manual review were added to the final evaluation dataset as shown in Tab. 2.

#### 4.3.2 High-precision Filter Combination

Using the final evaluation dataset with more comprehensive information on the relevance of retrieved trials, the impact of adding increasingly strict filters can be assessed. The results are shown in Tab. 5.

**Table 5.** Combination of different filters and impact on precision for the Hodgkin lymphoma and oesophageal cancer guideline update. There are no remaining results with unknown relevance for clinical trials after phase II.

| Search Queries | Retr. | | | | $\neg$ Retr. | | Metrics | | |
| --- | --- | --- | --- | --- | --- | --- | --- | --- | --- |
| (a) Oesophageal Cancer | Total | TP | ? | FP | TN | FN | Pr. | Re. | F<sub>1</sub> |
| All RCTs | 209 | 31 | 32 | 146 | 150 | 4 | 0.18 | **0.89** | 0.29 |
| ⊆ Phase ≥ II | 94 | 27 | **0** | 67 | 229 | 8 | 0.29 | 0.77 | 0.42 |
| ⊆ Phase ≥ III | 55 | 20 | 0 | 35 | 261 | 15 | 0.36 | 0.57 | **0.44** |
| ⊆ Excl. children | 55 | 20 | 0 | 35 | 261 | 15 | 0.36 | 0.57 | **0.44** |
| ⊆ Significant result | 37 | 15 | 0 | 22 | 274 | 20 | 0.41 | 0.43 | 0.42 |
| ⊆ $\exists$ Known interv. | 31 | 14 | 0 | 17 | 279 | 21 | **0.45** | 0.40 | 0.42 |
| ⊆ $\exists$ Unknown interv. | 28 | 12 | 0 | 16 | 280 | 23 | 0.43 | 0.34 | 0.38 |
| (b) Hodgkin Lymphoma |  |  |  |  |  |  |  |  |  |
| All RCTs | 80 | 40 | 25 | 15 | 20 | 0 | 0.73 | **1.00** | **0.84** |
| ⊆ Phase ≥ II | 45 | 32 | **0** | 13 | 22 | 8 | 0.71 | 0.80 | 0.75 |
| ⊆ Phase ≥ III | 28 | 24 | 0 | 4 | 31 | 16 | 0.86 | 0.60 | 0.71 |
| ⊆ Excl. children | 25 | 23 | 0 | 2 | 33 | 17 | 0.92 | 0.57 | 0.71 |
| ⊆ Significant result | 10 | 10 | 0 | 0 | 35 | 30 | **1.00** | 0.25 | 0.40 |
| ⊆ $\exists$ Known interv. | 10 | 10 | 0 | 0 | 35 | 30 | **1.00** | 0.25 | 0.40 |
| ⊆ $\exists$ Unknown interv. | 10 | 10 | 0 | 0 | 35 | 30 | **1.00** | 0.25 | 0.40 |

First, all retrieved results after filtering for clinical phases after phase II can be classified as either true positives (TP) or false positives (FP), i.e., there are no “?” entries after the second row for each data subset. Second, high levels of precision can be achieved by adding increasingly strict filtering criteria. Most gains in precision over the baseline (just filtering by populations) can already be achieved by selecting RCTs with phase III or later, resulting in +18pp. for oesophageal cancer and +13pp. for Hodgkin lymphoma. Excluding children does not affect the result set for oesophageal cancer, but improves precision by another 6pp. for Hodgkin lymphoma.

Adding a filter to retain only significant results achieves perfect precision on the Hodgkin lymphoma dataset: all results retrieved by the system are relevant for the guideline update, although with an overall recall of only 25.0 %. For oesophageal cancer, additional gains can be obtained by filtering for trials with at least one known intervention, i.e., one that is mentioned in the guideline, or at least one unknown intervention. Interestingly, the best precision for oesophageal cancer is achieved when considering only clinical trials, which have at least one *known* intervention. This could be attributed to the fact that a criterion for relevance of a trial is the right *comparison* arm, which is often equivalent to recommended clinical practice.

## 5 Discussion

The analysis of time lags has shown how important the right time point for updating a guideline can be. If not considered carefully, key results may be missed out in an update cycle. Building on that finding, we investigated how the NGE system can be used to retrieve potentially practice-changing signal publications at any given point during a guideline’s lifetime.

### 5.1 Implications

Our evaluation revealed that increased precision can be achieved by continuously adding stricter filtering criteria. Combining structured metadata, extracted population concepts, and information about the trial phase already provides comparatively high levels of precision. In both evaluation scenarios, precision could be further increased by classifying publications according to the statistical significance of their results. Including guideline context, i.e., which interventions are already recommended, can increase precision even further.

The given dataset represents a rather broad notion of relevance: not all publications that are reviewed need to be considered as update signals, and not all users of the NGE system will be interested in update signals. However, the results suggest that different combinations of filters provided by the system can increase precision for different user groups and their specific use-cases.

The relatively high number of results that were neither marked as included nor excluded in the provided screening dataset is rather surprising. This finding suggests the utility of our NGE system for quality-control in the systematic review processes, which has the goal of maximizing recall while ensuring reproducibility. For instance, if the system finds results not included in the original screening set due to the nature of the employed Boolean search string, this string can be adapted, e.g., to include currently not covered interventions.

### 5.2 Limitations

This section outlines the limitations of our work, highlighting potential challenges, constraints, and areas requiring further exploration.

#### 5.2.1 Limited Data for Time Lag Analysis

Our assessment of time lags in Section 4.2 was based on a comparatively small sample of guideline updates (22 new interventions), as only a relatively brief timespan for potential updates could be considered. As of now, no historical guideline versions are available through the GGPO CMS, which constrained the availability of data to those assembled after the first GGPOnc release in 2022 [20]. This small set of guidelines might not be fully representative of all update protocols encountered in practice. Although we found an overall time span of eight to ten years, which is consistent with suggestions from prior work, adding more data might provide a more reliable estimate for individual time lags.

Additional data could be gathered by considering guidelines from other medical specialties and countries. However, the analysis partially relies on a few curated metadata items (e.g., recommendation context) provided by the GGPO CMS, which might not be readily available for other guidelines or would at least require additional NLP components. Some guideline management tools such as the Magic app provide similar metadata, although it involves considerable manual curation efforts [54]. As more guideline organizations adopt international interoperability standards, like CPG-on-FHIR, our system can be enriched with more guideline sources [55]

#### 5.2.2 Metadata for Evidence Retrieval

Our NGE system relies on a few key metadata items for the retrieval of clinical trials, which imposes certain limitations. For instance, the recall with respect to the oesophageal cancer guideline update was lower than 100 %, as key population items were neither explicitly mentioned in the abstract of the publication nor in its assigned MeSH terms. This information might be recovered from the full-text of the publication, which also reflects how human experts screen literature for relevance. In prospective use, PubMed results are retrievable only after metadata for publication types is assigned. Although this process was recently automated by the NLM through the introduction of deep-learning based automated indexing using the MTIX system, a subset of publications is still subject to (slower) manual curation and quality-control [40, 58]. In practice, guideline expert panels may also rely on criteria for assessing relevancy that are different from the ones currently incorporated in the system. These could include bibliographic metrics and heuristics, like journal impact factors and authorship (e.g., well-known first or last authors).

#### 5.2.3 Focus on RCTs

Currently, our NGE system focuses on retrieving RCTs as the gold-standard for interventional study design because we expect that results of an individual RCT might have sufficient power to change the recommended clinical practice. In practice, guideline updates are often based on existing systematic reviews or meta-analyses of multiple RCTs, which provide an even higher level of evidence. These are currently not retrieved by the system, as key NLP components, such as the PICO tagger, were trained and evaluated on RCT abstracts only. Moreover, our interpretation of significant results might have to be broadened to incorporate non-inferiority trials. In contrast, lower-level evidence (observational studies, case reports) might be more relevant for fields where large RCTs are not the norm and may not even be practical to conduct. Other types of “gray” literature, such as meeting abstracts or government documents, could be included to obtain insights in a more timely manner [59]. This might account for the present risk of publication bias when considering published results only.

## 6 Conclusion

In this work, we described how NLP-derived metadata from reports of RCTs and clinical guidelines can be combined with structured resources, such as trial registries and precision oncology knowledge bases, to assemble an integrated database of medical evidence. Our NGE browser built upon this data was used to investigate time lags between publication of clinical research and recommendation as clinical practice. Observing that a considerable contributor to these time lags is the update frequency of guidelines due to lengthy evidence synthesis processes, we proposed an approach to identify signal publications through a targeted literature search within the NGE database. Our evaluation with respect to two guideline updates revealed that, through a combination of various selection criteria, large gains in precision can be achieved. The identification of additional and potentially relevant evidence beyond those included in the review highlights that the system is not only useful for guideline developers to identify update signals, but also for quality-control of existing search protocols. As an additional use case, clinical practitioners can use the system in conjunction with the latest guidelines to identify whether recommendations can still be considered up-to-date.

The system can be extended to import additional sources of primary and synthesized evidence to the database. A possible alternative to the free resources by the NLM are, for instance, the commercial database Embase, which provides access to the latest conference abstracts and richer search functionalities compared to PubMed [60]. Furthermore, the Cochrane Central Register of Controlled Trials (CENTRAL) is an alternative library of clinical trial reports, and includes data from PubMed/Medline, and Embase, but also the proprietary Cinahl database [61]. We can consider several additional registries for the primary registration of clinical trials, e.g., maintained by the European Union (Clinical Trials Information System / CTIS) or the BfArM (German Clinical Trials Register) [62]. The WHO operates the International Clinical Trials Registry Platform (ICTRP) combining data from multiple trial registries to provide a comprehensive global view of clinical trials. Alternatives to CIViC include OncoKB [63], My Cancer Genome [64], and Jax-Ckb [65]. However, integration of these data sources requires clearly documented access options, e.g., through APIs [26].

Additional guidelines could be integrated by considering other medical specialties, e.g., by applying the described NLP components to all other AWMF guidelines [66]. However, most guidelines currently do not provide fine-grained, recommendation-level metadata in a structured format, so they would need to be extracted from many heterogeneous documents, mostly in PDF format. Similarly, English-language guidelines, e.g., from the National Comprehensive Cancer Network (NCCN) or ASCO, could be easily integrated, as high-quality biomedical NER and NEN solutions for English texts are widely available [67].

## Acknowledgements

Parts of this work were generously supported by a grant from the German Federal Ministry of Research and Education (01ZZ2314N). Development of the Hodgkin lymphoma and oesophageal cancer guidelines was funded by the German Cancer Aid through the German Guideline Program in Oncology. The funders played no role in study design, data collection, analysis, and interpretation of data, or the writing of this manuscript. We thank Paul Freudenberger for his valuable input during the manual review of publications for the oesophageal cancer guideline.

## Data Availability

Recent versions of German oncology guidelines from the GGPO are available as GGPOnc releases on Zenodo: https://zenodo.org/records/12520623/.

Baseline and daily update dumps from PubMed can be downloaded directly from the NLM: https://pubmed.ncbi.nlm.nih.gov/download/.

Monthly dumps from ClinicalTrials.gov can be downloaded through the AACT project: https://aact.ctti-clinicaltrials.org/download/.

Nightly dumps from CIViC can be downloaded from the CIViC website: https://civicdb.org/releases/main/.

The literature screening datasets for the oesophageal cancer and Hodgkin lymphoma guideline updates can be made available upon request.

## Competing Interests

All authors declare no financial or non-financial competing interests.

## Author Contributions

FB conceptualized the work, performed the experiments, and wrote the initial draft of the manuscript. PW implemented the ETL processes and backend platform, and prepared the evaluation dataset. AO, NK, TK, TL, and NS created the guideline-related datasets, performed manual reviews of the system’s output, and contributed insight about guideline development processes. LW, MP, and BA supervised the project and were major contributors in writing the manuscript. All authors read and approved the final manuscript.

## Footnotes

1 We searched PubMed for “Clinical Trial”[Publication Type]

## Notes

### Competing Interest Statement

The authors have declared no competing interest.

## References

[1] Dave, T., Athaluri, S.A., Singh, S.: Chatgpt in medicine: an overview of its applications, advantages, limitations, future prospects, and ethical considerations. Frontiers in artificial intelligence 6, 1169595 (2023)

[2] Manion, F.J., Du, J., Wang, D., He, L., Lin, B., Wang, J., Wang, S., Eckels, D., Cervenka, J., Fiduccia, P.C., et al.: Accelerating evidence synthesis in observational studies: Development of a living natural language processing–assisted intelligent systematic literature review system. JMIR Medical Informatics 12(1), 54653 (2024)

[3] Brouwers, M.C., Florez, I.D., McNair, S.A., Vella, E.T., Yao, X.: Clinical practice guidelines: tools to support high quality patient care 49(a2), 145–152 (2019). Elsevier

[4] Shaitarova, A., Zaghir, J., Lavelli, A., Krauthammer, M., Rinaldi, F.: Exploring the latest highlights in medical natural language processing across multiple languages: A survey. Yearbook of Medical Informatics 32(1), 230 (2023)

[5] Hanney, S.R., Castle-Clarke, S., Grant, J., Guthrie, S., Henshall, C., Mestre-Ferrandiz, J., et al.: How long does biomedical research take? studying the time taken between biomedical and health research and its translation into products, policy, and practice. Health research policy and systems 13, 1–18 (2015)

[6] Subbiah, V.: The next generation of evidence-based medicine. Nature medicine 29(1), 49–58 (2023)

[7] Bastian, H., Glasziou, P., Chalmers, I.: Seventy-five trials and eleven systematic reviews a day: how will we ever keep up? PLoS med 7(9) (2010)

[8] Shekelle, P.G., Woolf, S.H., Eccles, M., Grimshaw, J.: Developing guidelines. Bmj 318(7183), 593–596 (1999)

[9] Schünemann, H.J., Wiercioch, W., Etxeandia, I., Falavigna, M., Santesso, N., Mustafa, R., Ventresca, M., Brignardello-Petersen, R., Laisaar, K.-T., Kowalski, S., et al.: Guidelines 2.0: systematic development of a comprehensive checklist for a successful guideline enterprise. Cmaj 186(3), 123–142 (2014)

[10] Karimi, S., Pohl, S., Scholer, F., Cavedon, L., Zobel, J.: Boolean versus ranked querying for biomedical systematic reviews. BMC medical informatics and decision making 10(1), 1–20 (2010)

[11] Straube, S., Heinz, J., Landsvogt, P., Friede, T.: Recall, precision, and coverage of literature searches in systematic reviews in occupational medicine: an overview of cochrane reviews. GMS Medizinische Informatik, Biometrie und Epidemiologie 17(1) (2021)

[12] Higgins, J.P., Thomas, J., Chandler, J., Cumpston, M., Li, T., Page, M.J., Welch, V.A. (eds.): Cochrane Handbook for Systematic Reviews of Interventions Version 6.5 (updated August 2024). Cochrane, Online (2024). https://www.training.cochrane.org/handbook [retrieved: Nov 1, 2024]

[13] Akl, E.A., Meerpohl, J.J., Elliott, J., et al.: Living systematic reviews: 4. living guideline recommendations. Journal of Clinical Epidemiology 91, 47–53 (2017)

[14] Elliott, J.H., Synnot, A., Turner, T., Simmonds, M., Akl, E.A., McDonald, S., et al.: Living systematic review: 1. introduction—the why, what, when, and how. Journal of clinical epidemiology 91, 23–30 (2017)

[15] Shekelle, P.G., Motala, A., Johnsen, B., Newberry, S.J.: Assessment of a method to detect signals for updating systematic reviews. Systematic reviews 3(1), 1–9 (2014)

[16] Somerfield, M.R., Bohlke, K., Browman, G.P., et al.: Innovations in American society of clinical oncology practice guideline development. Journal of Clinical Oncology 34(26), 3213–3220 (2016)

[17] Daly, M.E., Singh, N., Ismaila, N., Stage III NSCLC Guideline Expert Panel, M.: Management of stage III non–small cell lung cancer: ASCO rapid recommendation update. Journal of Clinical Oncology, 24 (2024)

[18] Lu, S., Kato, T., Dong, X., Ahn, M.-J., Quang, L.-V., Soparattanapaisarn, N., et al.: Osimertinib after chemoradiotherapy in stage III EGFR-mutated NSCLC. New England Journal of Medicine (2024)

[19] Bodenreider, O.: The Unified Medical Language System (UMLS): integrating biomedical terminology. Nucleic Acids Res. 32(Database issue), 267–70 (2004)

[20] Borchert, F., Lohr, C., Modersohn, L., Witt, J., Langer, T., Follmann, M., Gietzelt, M., Arnrich, B., Hahn, U., Schapranow, M.-P.: GGPONC 2.0 - the German clinical guideline corpus for oncology: Curation workflow, annotation policy, baseline NER taggers. In: Proceedings of the Language Resources and Evaluation Conference (LREC), pp. 3650–3660. European Language Resources Association, Marseille, France (2022)

[21] Kämmer, N., Borchert, F., Winkler, S., Melo, G., Schapranow, M.-P.: Resolving elliptical compounds in German medical text. In: The 22nd Workshop on Biomedical Natural Language Processing and BioNLP Shared Tasks, pp. 292–305. Association for Computational Linguistics, Toronto, Canada (2023)

[22] Borchert, F., Llorca, I., Roller, R., Arnrich, B., Schapranow, M.-P.: xMEN: A modular toolkit for cross-lingual medical entity normalization. arXiv [cs.CL] (Currently under review, pre-print available) 2310.11275 (2023)

[23] Marshall, I.J., Nye, B., Kuiper, J., Noel-Storr, A., Marshall, R., Maclean, R., et al.: Trialstreamer: A living, automatically updated database of clinical trial reports. Journal of the American Medical Informatics Association 27(12), 1903– 1912 (2020)

[24] (CTTI), C.T.T.I.: Aggregate Analysis of ClinicalTrials.gov (AACT) Database. https://aact.ctti-clinicaltrials.org/ [retrieved: Nov 1, 2024] (2024)

[25] Griffith, M., Spies, N.C., Krysiak, K., McMichael, J.F., Coffman, A.C., Danos, A.M., et al.: Civic is a community knowledgebase for expert crowdsourcing the clinical interpretation of variants in cancer. Nature genetics 49(2), 170 (2017)

[26] Borchert, F., Mock, A., Tomczak, A., Hügel, J., Alkarkoukly, S., Knurr, A., Volckmar, A.-L., Stenzinger, A., Schirmacher, P., Debus, J., Jäger, D., Longerich, T., Fröhling, S., Eils, R., Bougatf, N., Sax, U., Schapranow, M.-P.: Knowledge Bases and Software Support for Variant Interpretation in Precision Oncology. Briefings in Bioinformatics 22(6) (2021)

[27] Borchert, F., Meister, L., Langer, T., Follmann, M., Arnrich, B., Schapranow, M.-P.: Controversial trials first: Identifying disagreement between clinical guidelines and new evidence. In: AMIA Annual Symposium Proceedings, pp. 237–246. American Medical Informatics Association, San Diego, USA (2021)

[28] Balas, E.A.A., Boren, S.A.: Managing clinical knowledge for health care improvement. Yearbook of medical informatics 9(01), 65–70 (2000)

[29] Grant, J., Green, L., Mason, B.: Basic research and health: a reassessment of the scientific basis for the support of biomedical science. Research evaluation 12(3), 217–224 (2003)

[30] Morris, Z.S., Wooding, S., Grant, J.: The answer is 17 years, what is the question: understanding time lags in translational research. Journal of the royal society of medicine 104(12), 510–520 (2011)

[31] Pieper, D., Ober, P., Dressler, C., Schmidt, S., Mathes, T., Becker, M.: Effizientere Leitlinienerstellung–eine narrative übersichtsarbeit. Zeitschrift für Evidenz, Fortbildung und Qualität im Gesundheitswesen 146, 1–6 (2019)

[32] Alonso-Coello, P., Schünemann, H.J., Moberg, J., Brignardello-Petersen, R., Akl, E.A., Davoli, M., Treweek, S., Mustafa, R.A., Rada, G., Rosenbaum, S., et al.: Grade evidence to decision (etd) frameworks: a systematic and transparent approach to making well informed healthcare choices. 1: Introduction. bmj 353 (2016)

[33] Brouwers, M.C., Kho, M.E., Browman, G.P., Burgers, J.S., Cluzeau, F., Feder, G., Fervers, B., Graham, I.D., Grimshaw, J., Hanna, S.E., et al.: AGREE II: advancing guideline development, reporting and evaluation in health care. CMAJ 182(18), 839–842 (2010)

[34] Borah, R., Brown, A.W., Capers, P.L., Kaiser, K.A.: Analysis of the time and workers needed to conduct systematic reviews of medical interventions using data from the prospero registry. BMJ open 7(2), 012545 (2017)

[35] Shojania, K.G., Sampson, M., Ansari, M.T., Ji, J., Doucette, S., Moher, D.: How quickly do systematic reviews go out of date? a survival analysis. Annals of internal medicine 147(4), 224–233 (2007)

[36] Tsafnat, G., Glasziou, P., Choong, M.K., Dunn, A., Galgani, F., Coiera, E.: Systematic review automation technologies. Systematic reviews 3(1), 1–15 (2014)

[37] Van Dinter, R., Tekinerdogan, B., Catal, C.: Automation of systematic literature reviews: A systematic literature review. Information and Software Technology 136, 106589 (2021)

[38] Marshall, I.J., Kuiper, J., Wallace, B.C.: RobotReviewer: evaluation of a system for automatically assessing bias in clinical trials. Journal of the American Medical Informatics Association 23(1), 193–201 (2016)

[39] Hersh, W.: Information Retrieval: A Biomedical and Health Perspective, 4th edn. Health Informatics, vol. 4. Springer, Cham, Switzerland (2020)

[40] National Library of Medicine: MTIX: the Next-Generation Algorithm for Automated Indexing of Medline. NLM Tech Bull. 2024 Mar-Apr;(457):e4., https://www.nlm.nih.gov/pubs/techbull/ma24/ma24_mtix.html [retrieved: Nov 1, 2024] (2024)

[41] Fiorini, N., Canese, K., Starchenko, G., et al.: Best match: new relevance search for PubMed. PLoS biology 16(8) (2018)

[42] Trip Database Limited: Trip. http://www.tripdatabase.com/ [retrieved: Nov 1, 2024] (2020)

[43] Schapranow, M.-P., Kraus, M., Perscheid, C., Bock, C., Liedke, F., Plattner, H.: The medical knowledge cockpit: Real-time analysis of big medical data enabling precision medicine. In: 2015 IEEE International Conference on Bioinformatics and Biomedicine (BIBM), pp. 770–775 (2015). IEEE

[44] Faessler, E., Hahn, U.: Semedico: A comprehensive semantic search engine for the life sciences. In: Proceedings of ACL 2017, System Demonstrations, pp. 91–96. Association for Computational Linguistics, Vancouver, Canada (2017)

[45] Allot, A., Peng, Y., Wei, C.-H., Lee, K., Phan, L., Lu, Z.: Litvar: a semantic search engine for linking genomic variant data in pubmed and pmc. Nucleic acids research 46(W1), 530–536 (2018)

[46] Xue, L., Constant, N., Roberts, A., Kale, M., Al-Rfou, R., Siddhant, A., et al.: mT5: A massively multilingual pre-trained text-to-text transformer. In: Proceedings of the 2021 Conference of the North American Chapter of the Association for Computational Linguistics: Human Language Technologies, pp. 483–498. Association for Computational Linguistics, Online (2021)

[47] Bressem, K.K., Papaioannou, J.-M., Grundmann, P., Borchert, F., Adams, L.C., Liu, L., Busch, F., Xu, L., Loyen, J.P., Niehues, S.M., Augustin, M., Grosser, L., Makowski, M.R., Aerts, H.J.W.L., Löser, A.: medBERT.de: A comprehensive German BERT model for the medical domain. Expert Systems with Applications 237, 121598 (2024)

[48] Nye, B., Li, J.J., Patel, R., Yang, Y., Marshall, I., Nenkova, A., et al.: A corpus with multi-level annotations of patients, interventions and outcomes to support language processing for medical literature. In: Proceedings of the 56th Annual Meeting of the Association for Computational Linguistics (Volume 1: Long Papers), pp. 197–207. Association for Computational Linguistics, Melbourne, Australia (2018)

[49] Kanakarajan, K.r., Kundumani, B., Sankarasubbu, M.: BioELECTRA:pretrained biomedical text encoder using discriminators. In: Proceedings of the 20th Workshop on Biomedical Language Processing, pp. 143–154. Association for Computational Linguistics, Online (2021). Foo

[50] Neumann, M., King, D., Beltagy, I., Ammar, W.: ScispaCy: Fast and robust models for biomedical natural language processing. In: Proceedings of the 18th BioNLP Workshop and Shared Task, pp. 319–327. Association for Computational Linguistics, Florence, Italy (2019)

[51] Mohan, S., Li, D.: MedMentions: A large biomedical corpus annotated with UMLS concepts. In: Automated Knowledge Base Construction (AKBC) (2019)

[52] Singh, J.: Understanding ETL and Data Warehousing: Issues, Challenges and Importance. Lambert Academic Publishing, (2011)

[53] Seufferlein, T., Kopp, I., Post, S., Jonat, W., Kreienberg, R., Nothacker, M., et al.: Onkologische Leitlinien: Herausforderungen und zukünftige Entwicklungen. Forum 34, 277–283 (2019)

[54] Vandvik, P.O., Brandt, L., Alonso-Coello, P., Treweek, S., Akl, E.A., Kristiansen, A., et al.: Creating clinical practice guidelines we can trust, use, and share: a new era is imminent. Chest 144(2), 381–389 (2013)

[55] Lichtner, G., Alper, B.S., Jurth, C., Spies, C., Boeker, M., Meerpohl, J.J., von Dincklage, F.: Representation of evidence-based clinical practice guideline recommendations on fhir. Journal of Biomedical Informatics 139, 104305 (2023) 10.1016/j.jbi.2023.104305

[56] Gu, Y., Tinn, R., Cheng, H., Lucas, M., Usuyama, N., Liu, X., et al.: Domain-specific language model pretraining for biomedical natural language processing. ACM Transactions on Computing for Healthcare (HEALTH) 3(1), 1–23 (2021)

[57] DeYoung, J., Lehman, E., Nye, B., Marshall, I., Wallace, B.C.: Evidence inference 2.0: More data, better models. In: Proceedings of the 19th SIGBioMed Workshop on Biomedical Language Processing, pp. 123–132. Association for Computational Linguistics, Online (2020)

[58] Krithara, A., Mork, J.G., Nentidis, A., Paliouras, G.: The road from manual to automatic semantic indexing of biomedical literature: a 10 years journey. Frontiers in Research Metrics and Analytics 8, 1250930 (2023)

[59] Benzies, K.M., Premji, S., Hayden, K.A., Serrett, K.: State-of-the-evidence reviews: advantages and challenges of including grey literature. Worldviews on Evidence-Based Nursing 3(2), 55–61 (2006)

[60] Elsevier: Embase. https://www.elsevier.com/products/embase [retrieved: Nov 1 2024], 2024)

[61] Services, E.I.: CINAHL Database. https://www.ebsco.com/products/research-databases/cinahl-database [retrieved: Nov 1, 2024] (2024)

[62] Hasselblatt, H., Dreier, G., Antes, G., Schumacher, M.: The german clinical trials register: challenges and chances of implementing a bilingual registry. Journal of Evidence-Based Medicine 2(1), 36–40 (2009)

[63] Chakravarty, D., Gao, J., Phillips, S., Kundra, R., Zhang, H., Wang, J., et al.: Oncokb: A precision oncology knowledge base. JCO Precision Oncology 1, 1–16 (2017)

[64] Micheel, C.M., Lovly, C.M., Levy, M.A.: My Cancer Genome. Cancer Genetics 207(6), 289 (2014)

[65] Patterson, S.E., Liu, R., Statz, C.M., Durkin, D., Lakshminarayana, A., Mockus, S.M.: The clinical trial landscape in oncology and connectivity of somatic mutational profiles to targeted therapies. Human genomics 10, 4 (2016)

[66] Kopp, I., Encke, A., Lorenz, W.: Leitlinien als instrument der qualitätssicherung in der medizin: Das leitlinienprogramm der arbeitsgemeinschaft wissenschaftlicher medizinischer fachgesellschaften (awmf). Bundesgesundheitsblatt – Gesundheitsforschung – Gesundheitsschutz 45, 223–233 (2002)

[67] French, E., McInnes, B.T.: An overview of biomedical entity linking throughout the years. J. Biomed. Inform. 137, 104252 (2023)

