## Supplementary File 1 for "Next Generation Evidence: High-Precision Information Retrieval for Rapid Clinical Guideline Updates"

### Supplementary File 1: NLP and ETL Components

In this supplementary file, we share technical details on the data sources and employed ETL components, in particular those based on NLP models.

#### Clinical Guidelines (GGPO CMS)

The most recent versions of clinical guidelines are extracted from the CMS of the GGPO [1], as described by Borchert et al. [2]. The structured metadata (e.g., timestamps, versions, recommendation metadata, and literature references) need no further processing, and can be stored directly in the database according to their respective data types. In contrast, the textual guideline contents (recommendations and background texts) are used as inputs to an NLP pipeline for German medical documents. As the first step of the pipeline, compound noun phrases are resolved, such that entities within elliptical compounds can be more easily detected and normalized [3]. Second, NER tagging is applied using a model for long, fine-grained, nested entity spans, initialized from MEDBERT.DE [4]. Finally, the identified mentions are normalized to UMLS CUIs with XMEN [5]. The target KB for NEN is initialized from a subset of the UMLS containing the source vocabularies SNOMED CT, MESH, MEDDRA, and the NCI, and its German versions, if available. As the last step, a weakly supervised re-ranker is employed, i.e., a cross-encoder model trained on a machine-translated version of MEDMENTIONS [6].

The populations for each guideline are not explicitly defined with respect to any terminology system; a formal definition of a guideline's key questions according to PICO criteria is planned for future versions of the GGPO CMS. Therefore, I manually identify one or multiple CUIs as the root population of a guideline. For instance, the scope of the guideline for "prostate cancer" can be defined by the single concept "Prostatic Neoplasms" (CUI: C0033578), while the guideline for "Oro- and hypopharyngeal carcinoma" needs to be defined in terms of two concepts "Oropharyngeal Neoplasms" (C0029295) and "Hypopharyngeal Neoplasms" (C0020627). Upon import into the database, these root population CUIs are resolved to descendant concepts within the UMLS (exploiting the *narrower* (RN) / *child* (CD) relationships in the UMLS artifact `MRREL`), as shown in Fig. 1.

#### PubMed (RCT Reports)

As the source of RCT reports, we consider MEDLINE abstracts and their metadata, which are available as daily updated dumps from a file server maintained by the NLM. To detect RCTs among the downloaded MEDLINE articles, the NGE system relies on MEDLINE metadata for publication types and MESH terms. The included publication types are: "Randomized Controlled Trial", "Clinical Trial, Phase I", "Clinical Trial, Phase II", and "Clinical Trial, Phase III". Abstracts with the MESH term "Randomized Controlled Trials as Topic" are also included.

In the downloaded RCT abstracts, PICO spans are identified using a BIOELECTRA model, which was fine-tuned on the PICO span extraction task in the EBM-NLP corpus [7]. The pre-trained model from Kanakarajan et al. [8] is available on the HUGGING FACE Hub [9]. Within these PICO spans, all medical named entities are

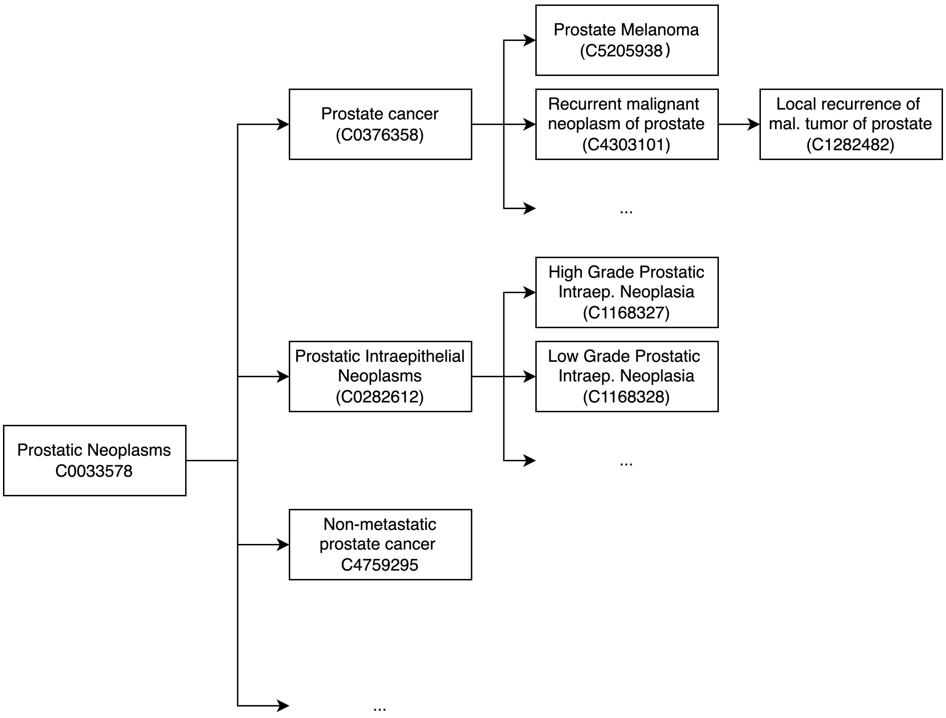


**Figure 1** Resolving top-level population concepts for a guideline topic to its descendants by the hierarchy modelled by the UMLS

identified using the SCISPACY NER pipeline (`en_core_sci_lg`). In addition, we rely on the integrated SCISPACY linker for NEN, which is equivalent to the TF-IDF linker in XMEN. However, the default SCISPACY linker is re-configured to use the same UMLS subset introduced for normalizing mentions in GGPONC. This way, all named entities are normalized to UMLS CUIs, and it can be inferred whether they refer to an intervention, population, or outcome, according to their overlap with PICO spans.

#### ClinicalTrials.gov (Registered Trials)

Information about a large number of (finished and ongoing) clinical trials is accessible through CLINICALTRIALS.GOV, a registry of clinical trials operated by the NLM. The data includes comprehensive information on eligibility and exclusion criteria, interventions, and study arms. For many trials, their results are also published in a structured format. Instead of accessing the data directly from CLINICALTRIALS.GOV, we rely on a tabular representation of the data provided through a monthly dump of the AACT database [10]. Interventions and conditions in AACT are already indexed with MESH terms, which can be easily mapped to UMLS CUIs through the UMLS metathesaurus. Moreover, the NLM maintains references between publications in MEDLINE and registered trials, which are incorporated into the database.

#### CIViC (Precision Oncology Knowledge Base)

We use assertions from the Clinical Interpretation of Variants in Cancer (CIViC) database, a crowdsourced community resource for the clinical actionability of cancer variants [11]. A recent survey [12] revealed that CIViC is among the most widely used KB in clinical practice with convenient access options, e.g., through nightly dumps of the full database content and an API. For data extraction, we use the Python library CIViCpy for downloading contents of the database [13]. Conceptually, the results are treated equivalently to clinical trials from PubMed; however, for results in CIViC, the system relies on curated rather than NLP-derived metadata.

### References


[1] Seufferlein, T., Kopp, I., Post, S., Jonat, W., Kreienberg, R., Nothacker, M., *et al.*: Onkologische Leitlinien: Herausforderungen und zukünftige Entwicklungen. Forum **34**, 277–283 (2019)

[2] Borchert, F., Lohr, C., Modersohn, L., Witt, J., Langer, T., Follmann, M., Gietzelt, M., Arnrich, B., Hahn, U., Schapranow, M.-P.: GGPONC 2.0 - the German clinical guideline corpus for oncology: Curation workflow, annotation policy, baseline NER taggers. In: Proceedings of the Language Resources and Evaluation Conference (LREC), pp. 3650–3660. European Language Resources Association, Marseille, France (2022)

[3] Kämmer, N., Borchert, F., Winkler, S., Melo, G., Schapranow, M.-P.: Resolving elliptical compounds in German medical text. In: The 22nd Workshop on Biomedical Natural Language Processing and BioNLP Shared Tasks, pp. 292–305. Association for Computational Linguistics, Toronto, Canada (2023)

[4] Bressem, K.K., Papaioannou, J.-M., Grundmann, P., Borchert, F., Adams, L.C., Liu, L., Busch, F., Xu, L., Loyen, J.P., Niehues, S.M., Augustin, M., Grosser, L., Makowski, M.R., Aerts, H.J.W.L., Löser, A.: medBERT.de: A comprehensive German BERT model for the medical domain. Expert Systems with Applications **237**, 121598 (2024)

[5] Borchert, F., Llorca, I., Roller, R., Arnrich, B., Schapranow, M.-P.: xMEN: A modular toolkit for cross-lingual medical entity normalization. arXiv [cs.CL] (Currently under review, pre-print available) **2310.11275** (2023)

[6] Mohan, S., Li, D.: MedMentions: A large biomedical corpus annotated with UMLS concepts. In: Automated Knowledge Base Construction (AKBC) (2019)

[7] Nye, B., Li, J.J., Patel, R., Yang, Y., Marshall, I., Nenkova, A., *et al.*: A corpus with multi-level annotations of patients, interventions and outcomes to support language processing for medical literature. In: Proceedings of the 56th Annual Meeting of the Association for Computational Linguistics (Volume 1: Long Papers), pp. 197–207. Association for Computational Linguistics, Melbourne,

Australia (2018)

[8] Kanakarajan, K.r., Kundumani, B., Sankarasubbu, M.: BioELECTRA:pretrained biomedical text encoder using discriminators. In: Proceedings of the 20th Workshop on Biomedical Language Processing, pp. 143–154. Association for Computational Linguistics, Online (2021). Foo

[9] Kanakarajan, K.R.: kamalkraj/BioELECTRA-PICO. https://huggingface.co/kamalkraj/BioELECTRA-PICO [retrieved: Nov 1, 2024] (2021)

[10] (CTTI), C.T.T.I.: Aggregate Analysis of ClinicalTrials.gov (AACT) Database. https://aact.ctti-clinicaltrials.org/ [retrieved: Nov 1, 2024] (2024)

[11] Griffith, M., Spies, N.C., Krysiak, K., McMichael, J.F., Coffman, A.C., Danos, A.M., *et al.*: Civic is a community knowledgebase for expert crowdsourcing the clinical interpretation of variants in cancer. Nature genetics **49**(2), 170 (2017)

[12] Borchert, F., Mock, A., Tomczak, A., Hügel, J., Alkarkoukly, S., Knurr, A., Volckmar, A.-L., Stenzinger, A., Schirmacher, P., Debus, J., Jäger, D., Longerich, T., Fröhling, S., Eils, R., Bougatf, N., Sax, U., Schapranow, M.-P.: Knowledge Bases and Software Support for Variant Interpretation in Precision Oncology. Briefings in Bioinformatics **22**(6) (2021)

[13] Wagner, A.H., Kiwala, S., Coffman, A.C., McMichael, J.F., Cotto, K.C., Mooney, T.B., *et al.*: Civicpy: a python software development and analysis toolkit for the civic knowledgebase. JCO Clinical Cancer Informatics **4**, 245–253 (2020)
