## Supplementary File 2 for "Next Generation Evidence: High-Precision Information Retrieval for Rapid Clinical Guideline Updates"

### Supplementary File 2: Features of the NGE Browser

This supplementary file provides additional details on the search functions of the NGE Browser, and how they are implemented. A screenshot from the criteria in the web interface is shown in Fig. 1.

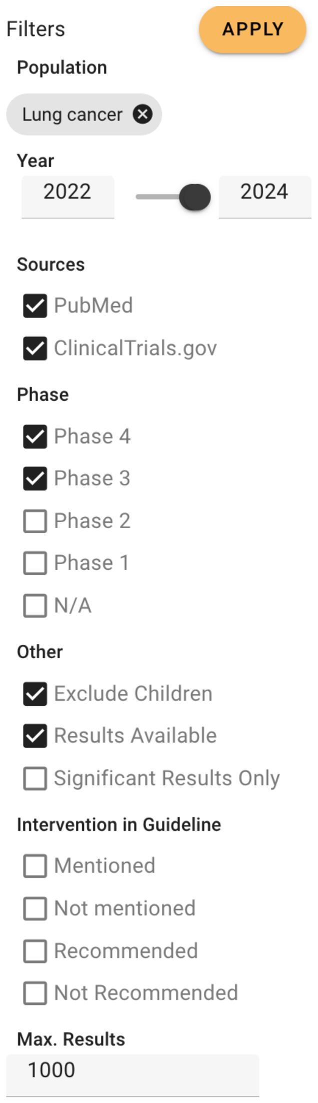


**Figure 1** A wrapped figure going nicely inside the text.

#### Population

The minimum input a user needs to provide is a population. Technically, this is a set of CUIs, from which at least one has to be present in the populations assigned to a clinical trial to be included in the result set. In the current user interface, the user can select a guideline topic as a proxy for a set of CUIs. For each topic, this set is pre-computed using the hierarchical expansion shown in Supplementary File 1.

#### Publication Date

For most use cases, the publication date of a trial is relevant, as recent results are usually more interesting than old ones. Moreover, for most guideline updates, there is a fixed date when the literature search has been completed. This date is essential for the evaluation of the system later in this chapter: in a retrospective setting, the result set should be limited to not include results past this date. In a prospective scenario, it is more desirable to find all evidence after this cutoff, which is potentially relevant for the guideline update, but could not be not covered by a previous literature screening. The default range for this filter is set to two years prior to the publication date of the selected guideline topic (minimum), and the current date (maximum).

#### Sources

Users can select the sources to be included in the result set, e.g., only include data from PubMed or ClinicalTrials.gov. Currently, only published, peer-reviewed reports of clinical trials are considered during guideline updates. However, for different surveillance strategies, ongoing clinical trials or results posted on ClinicalTrials.gov prior to a journal publication might be equally relevant.

#### Phase

Results can be filtered by the extracted trial phase; by default, only phase III and IV trials are returned, as these are the most relevant for the prospective scenario. Unfortunately, the phase is not unambiguously defined as metadata in the integrated sources. Therefore, a heuristic is applied to extract the trial phase, using a regular expression to extract numerical values in the following order: for PubMed articles, the system attempts to extract the phase from the publication title. If no phase can be extracted from the title, the assigned publication types and MeSH terms are considered. Finally, if no match is found, the abstract is considered. For ClinicalTrials.gov, there is a metadata item for the trial phase; if this is not set, the title and short description are considered as for PubMed articles. This heuristic has high recall, but might lead to some false positives, e.g., when an abstract mentions a prior trial of a lower phase. Many trials in oncology are combined phase I/II trials. For these reasons, a trial might have multiple phases assigned in the result set.

#### Interventions

Similar to the expansion of population CUIs shown in Supplementary File 1, matching of interventions in trials to guidelines also accounts for potential child–parent–relationships. Thus, an intervention is also considered present in a guideline, if any of its children are already mentioned. For instance, when a guideline mentions "Cetuximab" (C0995188), its parents "Monoclonal Antibodies" (C0003250) or "Protein Kinase Inhibitors" (C1449702) are also considered as already known.

#### Significant Results

For results from ClinicalTrials.gov, this information can be inferred from the structured results tab: any trial with a change in outcome associated with a $p$ value lower than 0.05 is considered significant. For published trial reports, this information needs to be obtained from the free-text abstract. To this end, we use a binary text classifier, which was trained by fine-tuning PubMedBERT [1] on a dataset derived from annotations in the Evidence Inference 2.0 dataset [2]. The classifier achieves an $F_1$ score of 0.84 for classifying trials reporting significant effects on the Evidence Inference test set (precision: 0.86, recall: 0.80).

#### Other Criteria

As most guidelines explicitly exclude childhood cancers from their scope, the search excludes trials related to children by default, i.e., by filtering by occurrence of the concept "Child" (C0008059) within populations (usually assigned as a MeSH term in PubMed). Moreover, only results from ClinicalTrials.gov with published results are included by default; when this filter is disabled, ongoing trials or completed trials without posted results are included as well, which tends to increase the result set substantially.

#### References

[1] Gu, Y., Tinn, R., Cheng, H., Lucas, M., Usuyama, N., Liu, X., *et al.*: Domain-specific language model pretraining for biomedical natural language processing. ACM Transactions on Computing for Healthcare (HEALTH) **3**(1), 1–23 (2021)

[2] DeYoung, J., Lehman, E., Nye, B., Marshall, I., Wallace, B.C.: Evidence inference 2.0: More data, better models. In: Proceedings of the 19th SIGBioMed Workshop on Biomedical Language Processing, pp. 123–132. Association for Computational Linguistics, Online (2020)
