## Supplementary File 3 for "Next Generation Evidence: High-Precision Information Retrieval for Rapid Clinical Guideline Updates"

### Supplementary File 3: Time Lag Analysis

This supplementary file provides additional details on the dataset used for analyzing time lags in translation of results from clinical trials to guidelines, and illustrates these time lags using representative examples.

#### New Interventions Across Guideline Updates

Tab. 1 shows the identified, newly recommended interventions extracted from oncology guideline updates in the time frame 2022–2024 (n=22).

| Guideline | Update | New Interventions |
|---|---|---|
| Endometrial cancer | 1.0 ▶ 2.0 | Trastuzumab, Dostarlimab, Pembrolizumab |
| Hepatocellular and biliary cancer | 3.0 ▶ 4.0 | Durvalumab, Tremelimumab |
| Lung cancer | 1.0 ▶ 2.0 | Durvalumab, Cemiplimab, Nivolumab + Ipilimumab, Amivantamab, Lorlatinib, Entrectinib, Repotrectinib, Larotrectinib, Selpercatinib, Pralsetinib, Capmatinib, Sotorasib |
| Oesophageal cancer | 3.0 ▶ 3.1 | Nivolumab, Pembrolizumab |
| Pancreatic cancer | 2.0 ▶ 3.0 | Low-molecular-weight heparin |
| Prostate cancer | 6.2 ▶ 7.0 | Niraparib, Talazoparib |

**Table 1** Newly recommended interventions per guideline within the considered timeframe

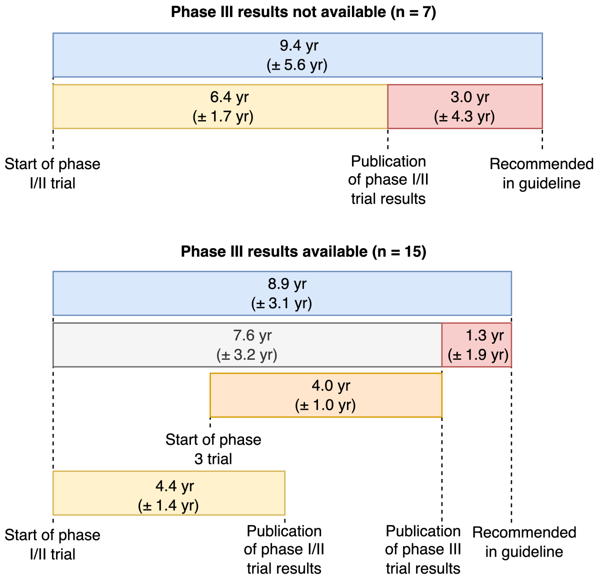


**Figure 1** Time span in years from the beginning of clinical trials until publication of their results and inclusion as a recommendation within a GGPO guideline (average and standard deviation)

#### Detailed Time Lags for Phase I/II vs. Phase III Trial Results

Using each of the combinations of CUI and guideline population in Tab. 1, the NGE database can be queried to identify all relevant trials that started prior to the publication of the respective updated guideline. We can now measure the time it took for each intervention from start to result publication of the first phase I/II and phase III trial, as well as the time between result publication and publication of the guideline update. Fig. 1 illustrates the average time spans and standard deviations for these combinations.

#### Qualitative Analysis

This section discusses selected examples of newly recommended interventions by using the timeline view of the NGE browser.

##### *Example: LMWH for Pancreatic Cancer*

Both the longest time lag for result publication of a phase II trial, and the difference between this publication and guideline recommendation refer to *low-molecular-weight heparins* (LMWH), a class of anticoagulant drugs, in the pancreatic cancer guideline. The timeline is shown in Fig. 2. It took almost nine years for the first trial on *Dalteparin*, a specific LMWH, for pancreatic cancer patients to be completed [1]. Additionally, it took another 12 years until anticoagulation was introduced as a topic in version 3.0 of the guideline.

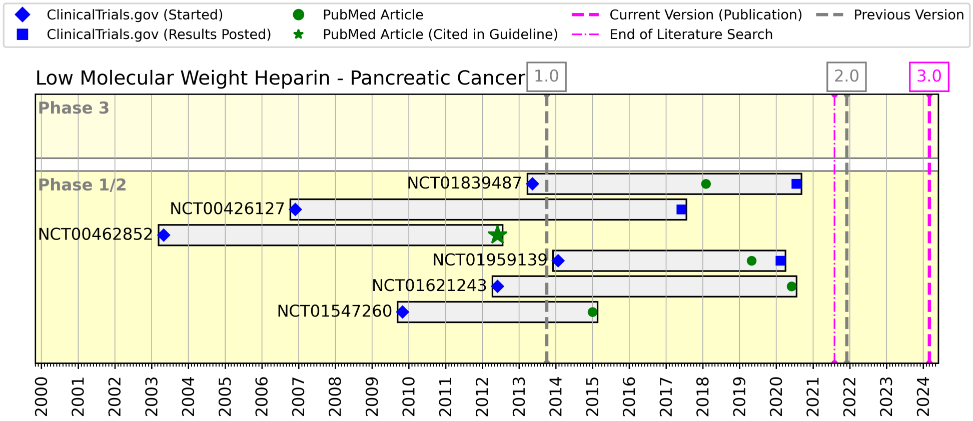


**Figure 2** Timeline of clinical trials concerning the intervention "Low Molecular Weight Heparin" (UMLS CUI: C0019139) for pancreatic cancer.

##### *Example: Amivantamab and Repotrectinib for Lung Cancer*

Two interesting examples concern the version 2.0 update of the lung cancer guideline. As shown in Fig. 3, *Amivantamab* was first recommended in version 2.0, based on early results of a phase I trial [2], while a phase III trial was still ongoing [3]. Similarly,

*Repotrectinib* was recommended based on an ongoing phase I/II trial (see Fig. 4); in this case, the guideline refers to a conference abstract (not included in the NGE database), as no peer-reviewed publication was available until very recently [4, 5].

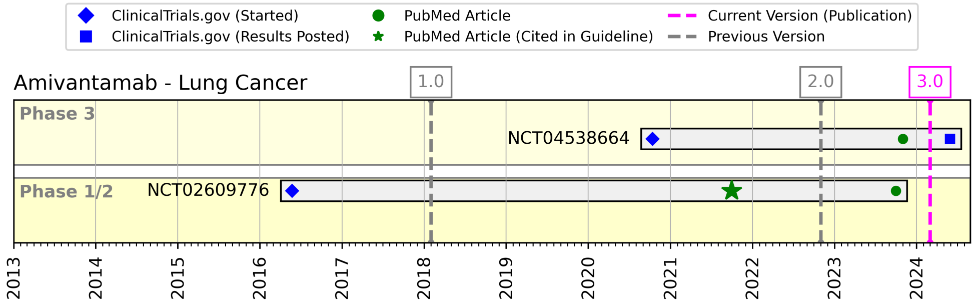


**Figure 3** Timeline of clinical trials investigating "Amivantamab" (C5446297) for lung cancer. Note that there was no specified date for literature search completion in version 3.0.

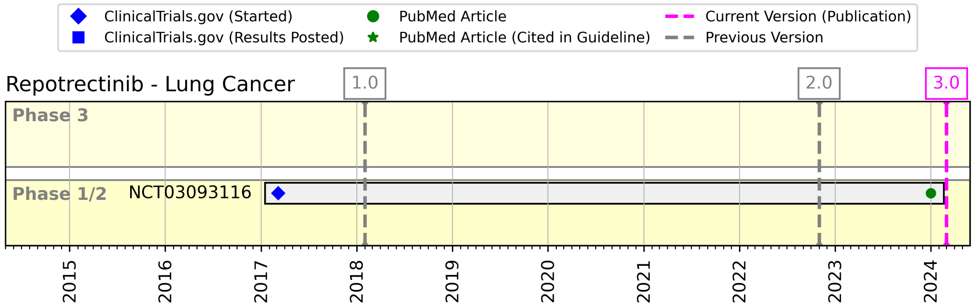


**Figure 4** Timeline of clinical trials investigating "Repotrectinib" (C4524909) for lung cancer.

##### *Example: Durvalumab for Lung Cancer*

Another example from the lung cancer guideline 2.0 update concerns *Durvalumab*, with a comparatively long timespan between completion of a phase III trial and recommendation in the guideline, as shown in Fig. 5. The recommendation was included in version 2.0, almost 5 years after results of the first phase III trial have been published. This can be explained by the long update interval between lung cancer version 1.0 and 2.0, which spanned around 4.5 years. The first *Durvalumab* phase III trial results [6] were published just before the final publication of the 1.0 release, and were thus not included in the systematic literature search for this version.

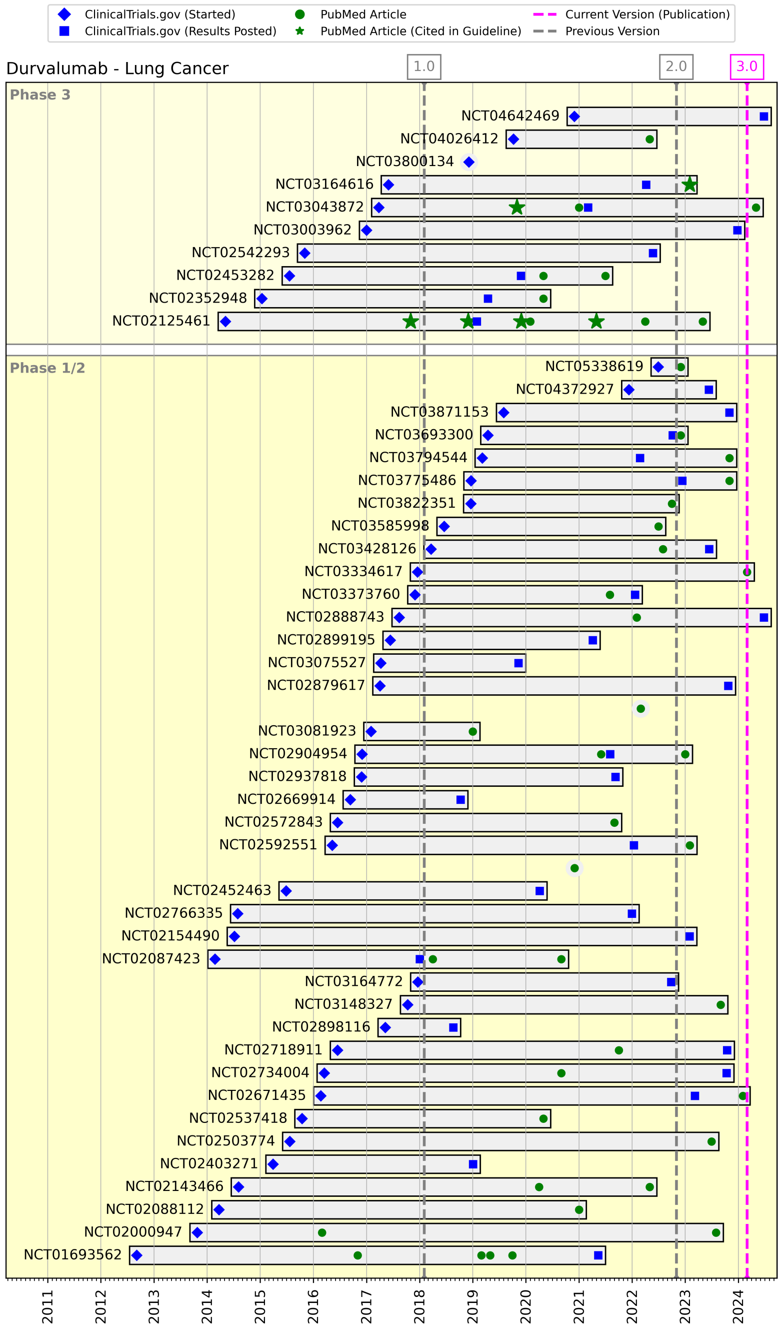


**Figure 5** Timeline of clinical trials investigating “Durvalumab” (C4055109) for lung cancer.

#### References


[1] Maraveyas, A., Waters, J., Roy, R., Fyfe, D., Propper, D., Lofts, F., *et al.*: Gemcitabine versus gemcitabine plus dalteparin thromboprophylaxis in pancreatic cancer. European journal of cancer **48**(9), 1283–1292 (2012)

[2] Park, K., Haura, E.B., Leighl, N.B., Mitchell, P., Shu, C.A., Girard, N., *et al.*: Amivantamab in egfr exon 20 insertion–mutated non–small-cell lung cancer progressing on platinum chemotherapy: initial results from the chrysalis phase i study. Journal of Clinical Oncology **39**(30), 3391–3402 (2021)

[3] Zhou, C., Tang, K.-J., Cho, B.C., Liu, B., Paz-Ares, L., Cheng, S., *et al.*: Amivantamab plus chemotherapy in nsclc with egfr exon 20 insertions. New England Journal of Medicine **389**(22), 2039–2051 (2023)

[4] Cho, B.C., Drilon, A.E., Doebele, R.C., Kim, D.-W., Lin, J.J., Lee, J., *et al.*: Safety and preliminary clinical activity of repotrectinib in patients with advanced ros1 fusion-positive non-small cell lung cancer (trident-1 study). J Clin Oncol **37**(15_suppl), 9011 (2019)

[5] Drilon, A., Camidge, D.R., Lin, J.J., Kim, S.-W., Solomon, B.J., Dziadziuszko, R., *et al.*: Repotrectinib in ros1 fusion–positive non–small-cell lung cancer. New England Journal of Medicine **390**(2), 118–131 (2024)

[6] Antonia, S.J., Villegas, A., Daniel, D., Vicente, D., Murakami, S., Hui, R., *et al.*: Durvalumab after chemoradiotherapy in stage iii non–small-cell lung cancer. New England Journal of Medicine **377**(20), 1919–1929 (2017)
